# Quality of life and anxiety in patients with non-muscle invasive bladder cancer: a prospective study

**DOI:** 10.1101/2020.08.13.20172973

**Authors:** Alexandros Vaioulis, Bonotis Konstantinos, Konstantinos Perivoliotis, Kiouvrekis Yiannis, Gravas Stavros, Tzortzis Vasilios, Karatzas Anastasios

## Abstract

**Introduction:** We evaluated anxiety and quality of life (QoL) in patients who were operated for non-muscle invasive bladder cancer (NMIBC)

**Methods:** The present study is a prospective analysis of patients with histopathologically confirmed NMIBCs after they were submitted to transurethral resection of the tumour (TURBT). Eligible were all adult patients with a single or multiple NMIBCs. All included patients followed therapy with either BCG or Epirubicin instillations. The SF-36 questionnaire Physical and Mental health aspects were used for QoL assessment. Similarly, the STAI-Y was introduced for the state (STAI-Y1) and trait anxiety (STAI-Y2) evaluation.

**Results:** In total, 117 eligible patients were included. Regarding SF-36 Physical a 6 months decrease was followed by an improvement at 12 months. Similarly, an increase of the SF-36 Mental health score was identified. In contrast to STAI-Y2, a long-term reduction of the state anxiety was identified. Preoperative SF-36 Physical was inversely correlated with age, while absence of alcohol was associated with lower mental health. Overall, patient characteristics, habits and the administered treatment did not affect the postoperative QoL and anxiety.

**Conclusions:** Patient QoL and anxiety improved during follow up. Although certain characteristics were related to QoL and anxiety, further larger scale studies are required.

## Introduction

Bladder cancer (BC) is one of the most common malignancies, accounting for almost 4.5% of all newly diagnosed cases worldwide (1). The annual incidence of BC is 180,000 new cases, whereas the mortality rate is estimated to be more than 55,000 patients per year. The gradually ageing population and the increased exposure to several risk factors are expected to further increment the burden of BC (2).

The majority of the new BC cases are diagnosed as non-muscle invasive BC (NMIBC) (3). Current approaches for the NMIBC management is based on the transurethral resection of the tumor followed by a strict follow-up schedule that may include intravesical administration of chemotherapeutic or immune-modifying regimens. Even though 5-year overall survival of NMIBC is 96%, recurrence is quite common, with the 1^st^ and the 5^th^ year rate being 33% and 50%, respectively. As a result, patients are required to undergo successive cystoscopies and adjuvant treatments (4).

Initial diagnosis, the strict surveillance program and the risk of recurrence have a major impact on the patients’ physical and mental health (5-7). Additionally, NIMBC intravesical treatment is associated with several quality of life (QoL) reducing adverse events, mainly lower urinary tract symptoms (LUTS) (8). Several series attempted to evaluate factors of the detrimental effect of the NMIBC therapeutic approaches on the psychological and physical health, with methodological issues (3, 5, 6, 9). The application of generic and widely validated tools, such as SF-36 and STAI-Y, allows for the overall assessment of the QoL and anxiety, respectively (10). However, current evidence regarding the perioperative QoL and anxiety patterns of NMIBC patients on the basis of SF-36 and STAI-Y are minimal (11).

In this study we tried to evaluate the QoL and the anxiety response in patients suffering from NMIBC and they were followed up thereafter.

## Methods

The present study is a prospective analysis of all patients submitted to surgery for NMIBC in our tertiary center, between 2018-2020. An institutional ethics approval was received. All eligible patients provided an informed consent. The study adhered to the Helsinki Declaration (12) and was reported on the basis of the STROBE guidelines (13).

Inclusion criteria were adult patients (age: 18-80 years) with histopathologically confirmed NMIBCs (pTis, pTa, pT1), following a scheduled intravesical therapy. Patients with severe comorbidities (ASA>III), muscular invasive bladder cancer, previous malignancy, recurrent bladder cancer, refusal to participate, and patients that were, either, not fluent Greek speakers, or who did not adhere to the follow up schedule were excluded.

All patients underwent endoscopic resection of the bladder tumour under spinal anesthesia. Three weeks postoperatively they initiated adjuvant intravesical therapy of either BCG or epirubicin instillations on an outpatient basis, as described elsewhere (14).

Demographics (age, gender, educational level, marital status, residence and employment status) and habitual data (smoking, alcohol) were recorded for all eligible patients. The SF-36 questionnaire was introduced for the QoL assessment. This tool has been previously validated in the Greek population (15) and its efficacy in the overall QoL evaluation has been extensively confirmed in various settings (16). All eight subscales of the questionnaire are sub-grouped in two major components, the Physical (Physical Functioning -PF, Role Physical-RP, Bodily Pain-BP and General Health-GH) and Mental (Vitality-VT, Mental Health-MH, Role Emotional-RE and Social Functioning-SF) health aspect. Results range from 0 to 100, with the latter being the optimal outcome. Consistently, anxiety levels were estimated by the validated State-Trait Anxiety Inventory (STAI-Y) questionnaire (17). The two subscales of this tool address the state (STAI-Y1) and trait anxiety (STAI-Y2) of the subject. Scores ranging from 29-39 were considered as low, whereas values greater that 60 were translated as increased anxiety. Patient evaluation was performed 2 weeks preoperatively and during the scheduled postoperative follow up (3, 6 and 12 months after surgery) examination. All data was secured in an encrypted electronic repository.

The primary endpoint of our study was the appraisal of the SF-36 Physical health aspect variations during the follow up period. Secondary endpoints included the evaluation of the time fluctuations of SF-36 Mental, STAI-Y1 and STAI-Y2. Moreover, an association analysis of patient characteristics with the preoperative and postoperative QoL and anxiety measurements was performed. Based on the reliability of the questionnaires, the minimum required number of sampling observations (power: 80%, effect size: 0.3, type I error: 5%) was estimated at the level of 90 cases.

All data underwent a Shapiro-Wilk test for normality. Independent samples t test was used for continuous variables. Given an incompetence to confirm normality, the evaluation of a possible relationship between distinct patient characteristics and the preoperative QoL and anxiety levels was based on the Mann-Whitney U and the Kruskal-Wallis H test. In order to examine the time-related variations of the questionnaire measurements, a repeated measures ANOVA model (RM-ANOVA) was introduced. Violation of the sphericity assumption was assessed by the Mauchly’s test of sphericity. If a violation was confirmed, then a Greenhouse-Geisser correction was applied. Post hoc comparisons and interactions of time endpoints and patient characteristics were calculated. The effect of distinct characteristics on the measured variables was estimated by the F test. Cronbach’s Alpha was, also, calculated for the evaluation of the internal consistency of the questionnaires in each time endpoint.

Categorical variables were provided as N (Percentage), whereas continuous data were reported as Mean (Standard Deviation). For all analyses, mean ± 2SE was displayed through all time endpoints. Statistical significance was considered at the level of p<0.05. All analyses were completed in SPSS version 23 software (SPSS Inc. Chicago, IL, USA).

## Results

In total, 117 eligible patients were identified during the enrollment period. Table 1 summarizes the demographic characteristics. 97 males and 11 females were included. Most cases (63%) were >66 years old. The majority of the patients had completed the primary education (45.4%) and were married (85.2%). Patient allocation in terms of residence and employment status is also provided in Table 1. Regarding habits, a higher rate of cases was never exposed to alcohol (35.2%), in contrast to smoking (11.1%).

**Table 1:** Patients’ demographics

|  |  | N | Percentage (%) |
| --- | --- | --- | --- |
| Gender | Male | 97 | 89.8 |
|  | Female | 11 | 10.2 |
| Age | <66 years | 40 | 37.0 |
|  | 66+ years | 68 | 63.0 |
| Educational Level | Illiterate | 3 | 2.8 |
|  | Primary Education | 49 | 45.4 |
|  | Secondary Education | 26 | 24.1 |
|  | Higher Education | 26 | 24.1 |
|  | MSc | 4 | 3.7 |
|  | PhD | 0 | 0.0 |
| Marital Status | Unmarried | 2 | 1.9 |
|  | Married | 92 | 85.2 |
|  | Divorced | 8 | 7.4 |
|  | Widowed | 4 | 3.7 |
|  | Cohabitation | 2 | 1.9 |
| Residence | Local Prefecture | 75 | 69.4 |
|  | Local Region | 27 | 25.0 |
|  | Outside Local Region | 6 | 5.6 |
| Employment Status | Employed | 20 | 18.5 |
|  | Retired | 83 | 76.9 |
|  | Unemployed | 4 | 3.7 |
|  | Household | 1 | 0.9 |
| Smoking | No | 12 | 11.1 |
|  | Ex-smoker | 60 | 55.6 |
|  | Yes | 36 | 33.3 |
| Alcohol | No | 38 | 35.2 |
|  | Social consumption | 56 | 51.9 |
|  | Daily | 14 | 13.0 |
| Treatment | BCG | 91 | 84.3 |
|  | Epirubicin | 17 | 15.7 |

The fluctuation of the SF-36 Physical over the progressive time endpoints is visualized in Figure 1. Mean values at 3 months were comparable to the preoperative measurements. The 6 months decrease was followed by a significant improvement at 12 months. Overall, RM-ANOVA (Table 2) confirmed the presence of a significant time variation (p=0.008).

**Figure 1.**
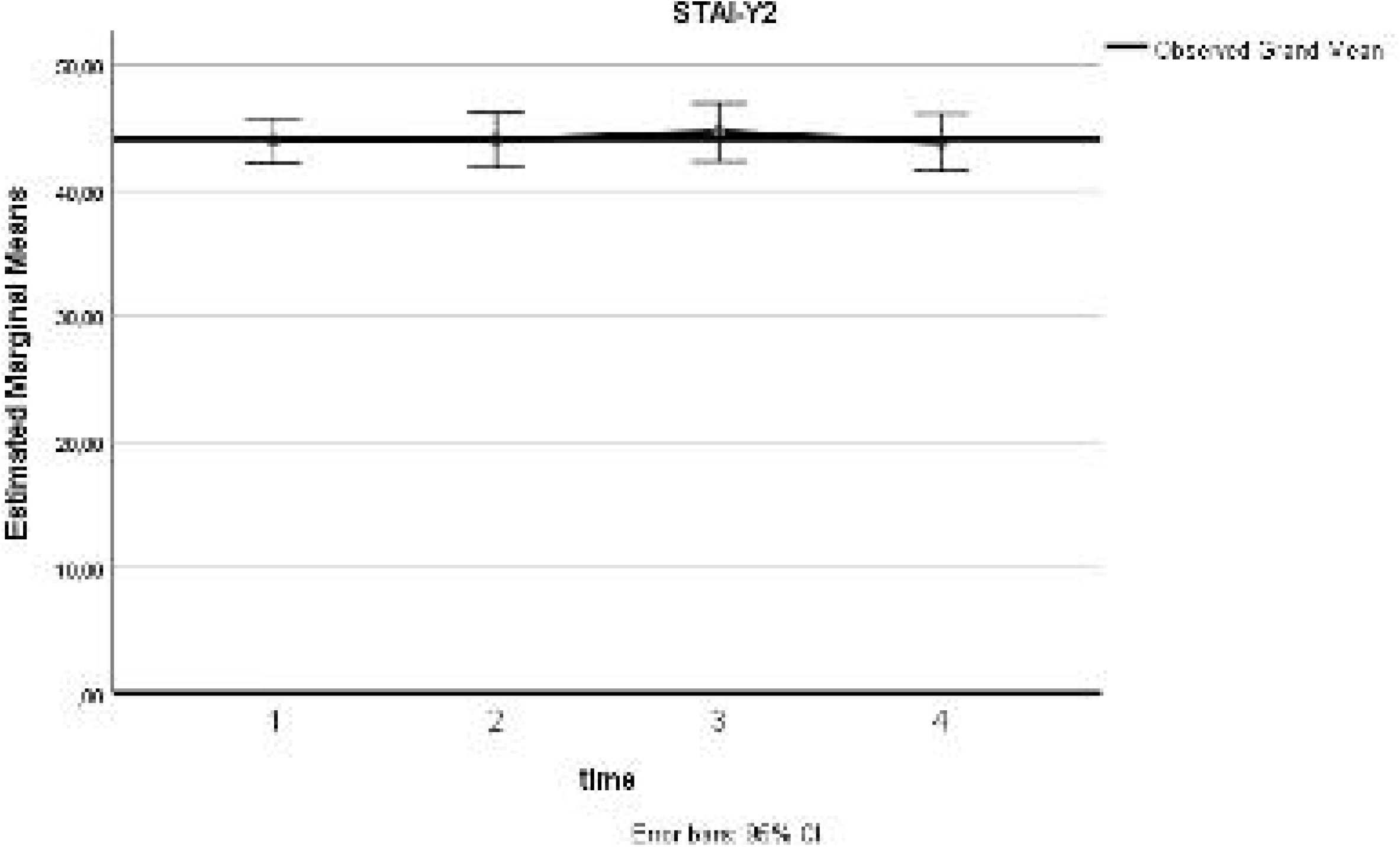
SF-36 Physical

**Table 2:** Basic questionnaire results

| Subscale | Time | Mean | Standard<br>Deviation | Mauchly's<br>Sphericity P | Time<br>Variation P |
| --- | --- | --- | --- | --- | --- |
| SF-36 Physical | 2 weeks preoperatively | 64.6 | 24.1 | **0.011** | **0.008\*** |
|  | 3 months postoperatively | 64.3 | 24.9 |  |  |
|  | 6 months postoperatively | 60.0 | 25.9 |  |  |
|  | 12 months postoperatively | 71.0 | 19.4 |  |  |
| SF-36 Mental | 2 weeks preoperatively | 64.2 | 24.4 | **0.011** | **0.03\*** |
|  | 3 months postoperatively | 61.1 | 26.2 |  |  |
|  | 6 months postoperatively | 64.1 | 22.7 |  |  |
|  | 12 months postoperatively | 72.2 | 18.3 |  |  |
| STAI-Y1 | 2 weeks preoperatively | 41.2 | 11.1 | **0.023** | **0.001\*** |
|  | 3 months postoperatively | 42.1 | 14.7 |  |  |
|  | 6 months postoperatively | 40.7 | 13.6 |  |  |
|  | 12 months postoperatively | 33.0 | 9.3 |  |  |
| STAI-Y2 | 2 weeks preoperatively | 44.0 | 9.3 | 0.362 | 0.945 |
|  | 3 months postoperatively | 44.0 | 11.3 |  |  |
|  | 6 months postoperatively | 44.6 | 12.1 |  |  |
|  | 12 months postoperatively | 43.8 | 11.8 |  |  |

Similarly, a significant (p=0.03) increase of the SF-36 Mental health score (Figure 2) was identified. As shown in Table 2, a higher 12-month mean value was estimated (72.2), compared to the respective preoperative assessment (64.2). In terms of anxiety, analysis confirmed a long-term decrease (Figure 3) of the state anxiety (preoperative STAI-Y1: 41.2, 12 months STAI-Y2: 33, p=0.001). In contrast to these, STAI-Y2 reports (Figure 4) were consistent over the time endpoints (p=0.945).

**Figure 2.**
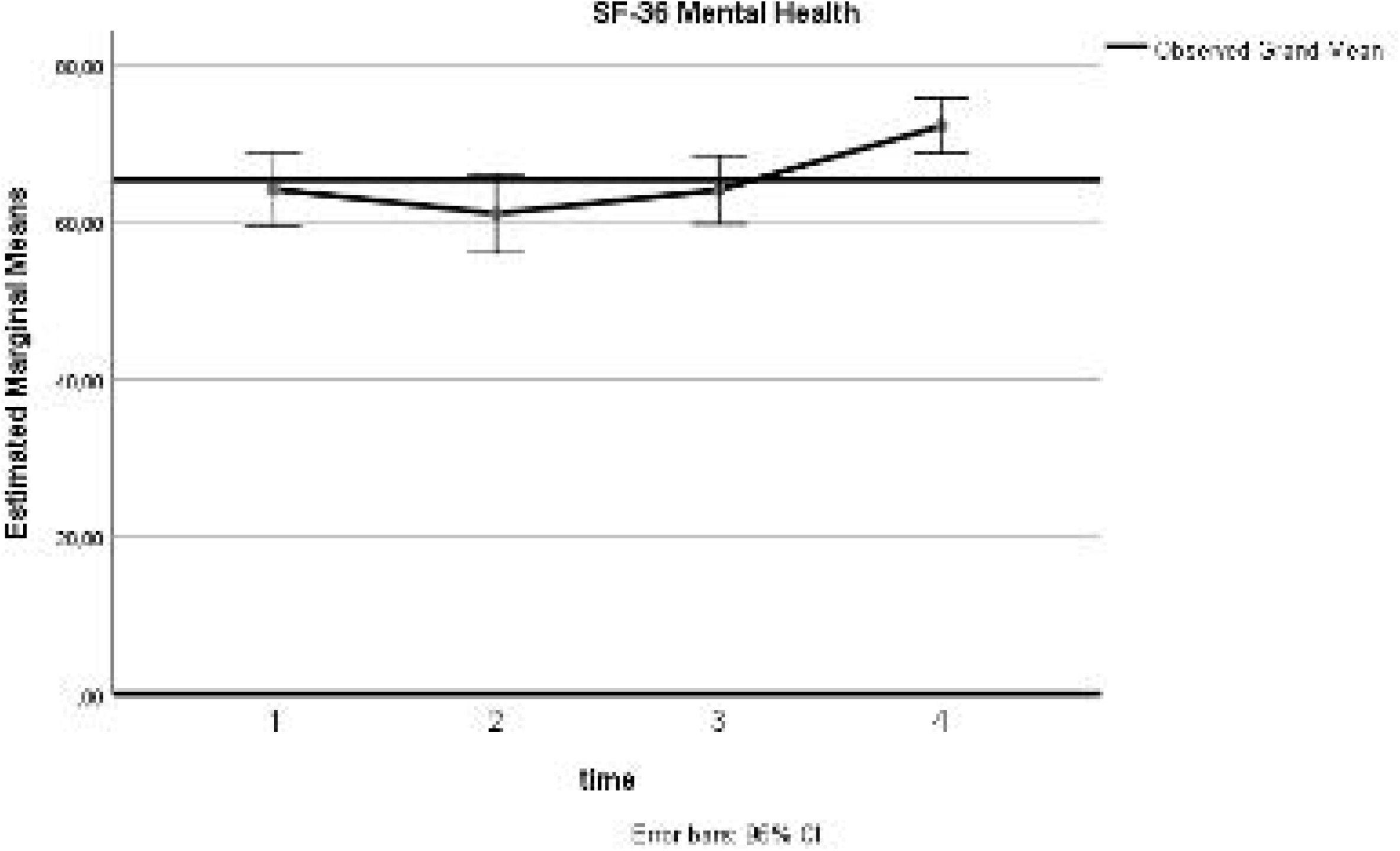
SF-36 Mental

**Figure 3.**
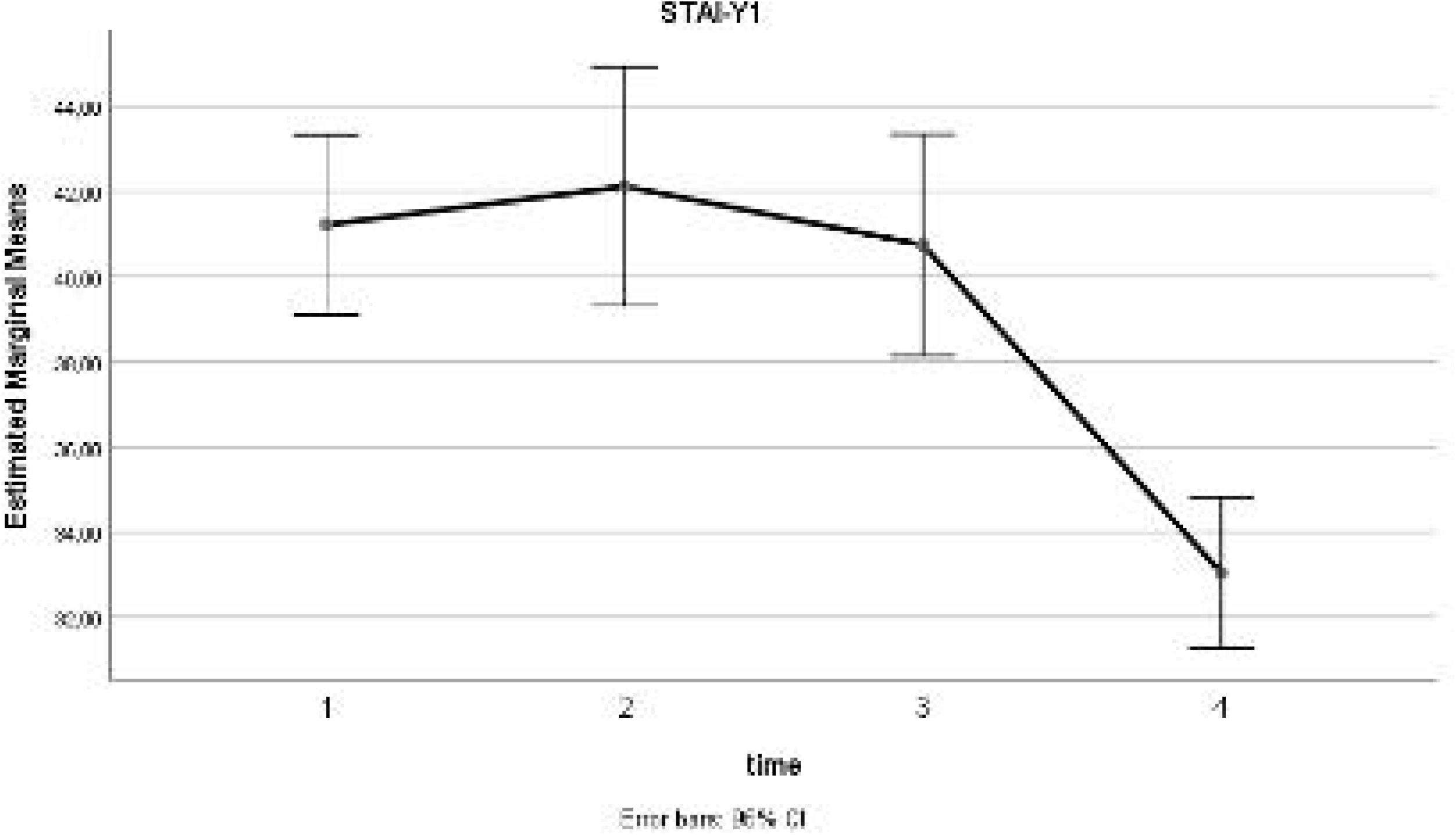
STAI-Y1

**Figure 4.**
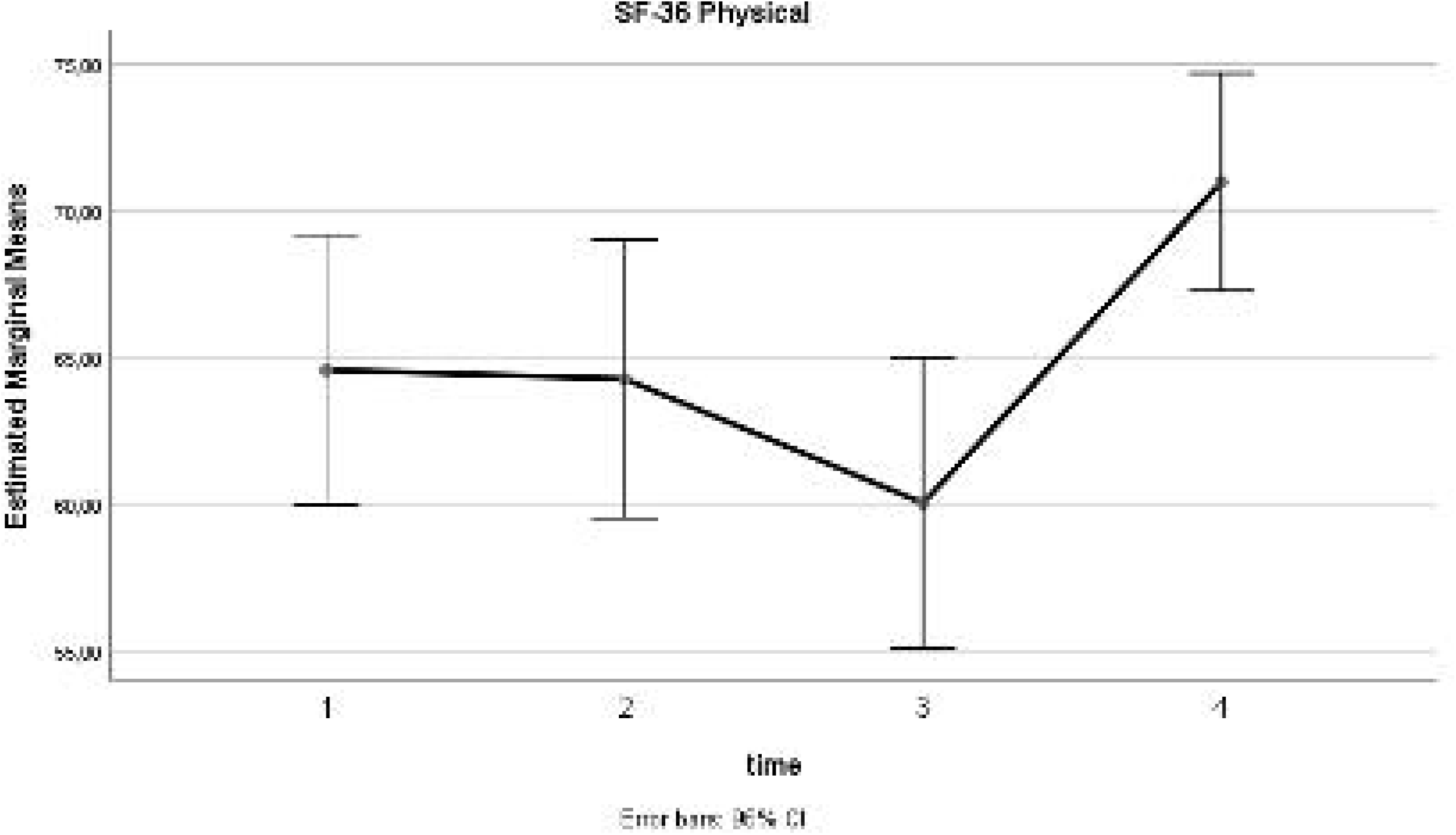
STAI-Y

Preoperative SF-36 Physical scores were inversely (p=0.029) correlated with age (Table 3 and Supplementary Material Figures). Absence of alcohol consumption was associated with significantly lower values in mental health (SF-36 Mental p=0.003). Gender, educational level and smoking did not affect the preoperative QoL and anxiety levels (Table 4 and Supplementary Material Figures).

**Table 3.** The effect of patient characteristics on preoperative questionnaire results.

| Subscale | Factor | P |
| --- | --- | --- |
| SF-36 Physical | Gender | 0.499 |
|  | **Age** | **0.029**<br>**(Z=-2.179)** |
|  | Educational Level | 0.188 |
|  | Marital Status | 0.475 |
|  | Residence | 0.123 |
|  | Smoking | 0.862 |
|  | Alcohol | 0.325 |
| SF-36 Mental | Gender | 0.733 |
|  | Age | 0.483 |
|  | Educational Level | 0.386 |
|  | Marital Status | 0.413 |
|  | Residence | 0.878 |
|  | Smoking | 0.424 |
|  | **Alcohol** | **0.003**<br>**($\chi^2(2)=11.492$)** |
| STAI-Y1 | Gender | 0.764 |
|  | Age | 0.597 |
|  | Educational Level | 0.299 |
|  | Marital Status | 0.361 |
|  | Residence | **0.040** |
|  | Smoking | 0.313 |
|  | Alcohol | 0.109 |
| STAI-Y2 | Gender | 0.452 |
|  | Age | 0.386 |
|  | Educational Level | 0.495 |
|  | Marital Status | 0.170 |
|  | Residence | 0.213 |
|  | Smoking | 0.575 |
|  | Alcohol | 0.102 |

**Table 4.** Correlation of patient characteristics with quality of life and anxiety measurements.

| Subscale | Factor | Mauchly's Sphericity P | Interaction between Factor and Time P | Overall Effect of Factor P |
| --- | --- | --- | --- | --- |
| SF-36 Physical | Treatment | **0.013** | 0.216* | 0.428 |
|  | Gender | **0.013** | 0.558* | 0.776 |
|  | Age | **0.002** | **0.011\*** | 0.605 |
|  | Educational Level | **0.012** | 0.222* | 0.863 |
|  | Marital Status | **0.017** | 0.754* | 0.085 |
|  | Residence | **0.011** | 0.479* | 0.529 |
|  | Smoking | **0.003** | 0.108* | 0.419 |
|  | Alcohol | **0.012** | **0.015\*** | 0.89 |
| SF-36 Mental | Treatment | **0.023** | 0.06* | 0.83 |
|  | Gender | **0.009** | 0.265* | 0.379 |
|  | Age | **0.012** | 0.241* | 0.822 |
|  | Educational Level | **0.022** | 0.237* | 0.785 |
|  | Marital Status | **0.021** | 0.502* | 0.203 |
|  | Residence | **0.006** | 0.291* | 0.455 |
|  | Smoking | **0.007** | 0.816* | 0.641 |
|  | Alcohol | **0.006** | **0.02\*** | 0.156 |
| STAI-Y1 | Treatment | 0.051 | 0.059 | 0.253 |
|  | Gender | **0.016** | 0.4* | 0.872 |
|  | Age | **0.025** | 0.84* | 0.307 |
|  | Educational Level | **0.043** | 0.811* | 0.60 |
|  | Marital Status | **0.025** | 0.758* | 0.962 |
|  | Residence | **0.006** | 0.117* | 0.672 |
|  | Smoking | **0.027** | 0.802* | 0.707 |
|  | Alcohol | **0.017** | **0.011\*** | 0.071 |
| STAI-Y2 | Treatment | 0.305 | 0.157 | 0.798 |
|  | Gender | 0.341 | 0.71 | 0.809 |
|  | Age | 0.398 | 0.446 | 0.22 |
|  | Educational Level | 0.398 | 0.229 | 0.763 |
|  | Marital Status | 0.301 | 0.799 | 0.596 |
|  | Residence | 0.291 | 0.377 | 0.346 |
|  | Smoking | 0.516 | 0.202 | 0.828 |
|  | Alcohol | 0.179 | **0.003** | 0.198 |

A significant interaction of age (p=0.011) and alcohol consumption (p=0.015) with time in the Physical aspect of the SF-36 was documented (Supplementary Material Figures). However, there was no significant effect (p=0.605 and p=0.89) of these variables on the mean tool measurements. Moreover, a significantly different pattern of SF-36 Mental (p=0.02), STAI-Y1 (p=0.011) and STAI-Y2 (p=0.003) scores, over the successive time endpoints was identified in the various alcohol consumption subgroups. A significant effect of alcohol was not confirmed in any tool. Overall, patient characteristics, habits and the administered treatment did not affect the postoperative QoL and anxiety.

Based on the sample size, the power of the present study was estimated at the level of 87.1%. Post-hoc analysis of the questionnaires’ internal consistency revealed an adequate level of validity (Cronbach Alpha: 72.4%-93.7%) (Table 5). Exception to these were the 6-months SF-36 Mental and the 12-months STAI-Y1 measurements (6% and 35% respectively).

**Table 5.** Internal consistency of the questionnaires’ subscales using Cronbach Alpha

| Subscale | Time | Cronbach Alpha (%) |
| --- | --- | --- |
| SF-36 Physical | 2 weeks preoperatively | 72.4 |
|  | 3 months postoperatively | 78.8 |
|  | 6 months postoperatively | 80.2 |
|  | 12 months postoperatively | 81.7 |
| SF-36 Mental | 2 weeks preoperatively | 83.1 |
|  | 3 months postoperatively | 86.4 |
|  | 6 months postoperatively | 6.0 |
|  | 12 months postoperatively | 78.7 |
| STAI-Y1 | 2 weeks preoperatively | 88.6 |
|  | 3 months postoperatively | 93.7 |
|  | 6 months postoperatively | 92.8 |
|  | 12 months postoperatively | 35.0 |
| STAI-Y2 | 2 weeks preoperatively | 80.1 |
|  | 3 months postoperatively | 86.6 |
|  | 6 months postoperatively | 87.0 |
|  | 12 months postoperatively | 84.8 |

## Discussion

### Summary of evidence

BC incidence ranges from 3.3 to 9.5/ 100,000 new cases per year. Several genetic and habitual risk factors have been related to the increased BC rate. The most prominent among them are hereditary polymorphisms, occupational exposure, non-fiber dietary habits, smoking and alcohol consumption (18). Regarding NMIBC, adjuvant maintenance therapy in various protocols with chemotherapeutic or immune-modifying agents is deemed as necessary in most cases (4). Currently, the optimal duration and frequency of the intravesical treatment is debatable (19). Among the various regimens, BCG displays a significantly higher efficacy in recurrence risk reduction, at the cost of increased morbidity, e.g. hematuria, orchitis, cystitis, fever and sepsis (20). Besides these commonly reported adverse events, the impact of the initial diagnosis and overall management of the NMIBC patients on the QoL plays important role and has been recently addressed in several studies (3).

QoL corresponds to the fulfillment level of an individual’s physical, mental, and social needs and therefore is attained through the interaction of various components. These QoL affecting factors include the somatic and emotional health, the interpersonal relationships, and the overall living standard. The importance of the latter is highlighted by the significant impact of the financial, employment and educational status on QoL. Besides biological and psychological health, health related QoL (HRQoL) involves the assessment of the functionality and the well-being of an individual. Therefore, the interference of personal expectations and health-related perceptions result to a subjective evaluation of HRQoL (21).

The inherent difficulty in differentiating between the subjective and objective components alongside the presence of several confounding factors, renders QoL assessment a challenging task (22). Taking into consideration the variability of the available techniques, valid QoL evaluation is based on the suitability of the applied methods. Although the design of most of these tools addressed samples with heterogeneous base characteristics, the QoL appraisal in specific patient categories required the introduction of specialized questionnaires. Overall, QoL assessment tools are allocated in two subgroups. General indicators study generic aspects of physical and psycho-social functionality, in contrast to the specific assessment tools where emphasis is placed on the individual’s perception regarding health and HRQoL, in distinct patient sub-categories (23).

Besides a considerable economic cost, NMIBC has a significant impact on a patient’s QoL. The initial cancer diagnosis, the frequent endoscopies and the adverse events related to the intravesical therapy affect the physical function, the mental health, the reported fatigue and the role function (5). The detrimental effect of a BC diagnosis on QoL was reported by Singer et al. (24). Similarly, Fung et al. confirmed a significant post-diagnosis reduction of the physical and mental scores (25). It was also noted that the effect on the physical domain had a duration of 10 years. A NMIBC related poor emotional function has been, also, validated. In a prospective analysis by Schmidt et al., it was estimated that the impact of NMIBC on mental health was detectable during the 6 month follow-up period (26). However, in case of multiple TURBs, it was found that the initial impairment on mental health, gradually ameliorated. In terms of emotional role and physical and social functioning, a minimization of the estimated scores is expected at the 2^nd^ or 3^rd^ reoperation (16).

In our study, SF-36 physical health scores reached a nadir at 6 months postoperatively, whereas the mental component displayed a minimum value at 3 months. A gradual improvement, that even reached higher values compared to the preoperative scores, over the remaining follow-up period was identified in both tools. However, these findings are in contrast with the respective literature reports. In the BOXIT trial, patients with no NMIBC progression or recurrence reached a minimal EQ-5D score at 2 months postoperatively, with no further improvement (27). Similarly data from a multicenter prospective cohort trial reported a plateau in the SF-36 mental and physical scores throughout the study period (26). Our data, also, suggested a significant association of age and alcohol with the Physical and Mental health, respectively. Similarly, in a study of 1160 BCs, Yu et al., reported an improved physical component in young and healthy, single males, that did not smoke and were diagnosed with a NMIBC. Moreover, gender, age, tumor stage and comorbidity status, also, affected preoperative mental health (3). Although various patient characteristics interacted with time QoL measurements, none of them was an independent QoL affecting factor. Evidence from recent studies, though, suggest that postoperative HRQoL is significantly associated with modifiable behaviors such as physical activity, dietary habits and smoking (6).

Furthermore, anxiety levels are an important determinant of the treatment efficiency. Anxiety is defined as the unpleasant sense of fear in an unknown and ill-defined threat, followed by a variety of emotional and hemodynamic manifestations. Assessment of anxiety is based on the evaluation of both state and trait anxiety levels (17). The former is regarded as a temporary condition that can re-emerge under the proper stimulus, whereas the latter represents the specific and repeatable way that an individual reacts or perceives certain conditions (28). Preoperative anxiety is a well-documented entity with an almost 80% incidence (29). The physical and emotional distress symptoms have a wide severity range and can present several weeks preoperatively (28). Furthermore, increased perioperative anxiety is related to suboptimal patient recovery, impaired immunologic function and increased morbidity and mortality. Despite initial diagnosis, socioeconomic factors, such as educational and marital status were predictive factors of preoperative anxiety. Preoperative distress, combined with postoperative pain and smoking are considered as independent postoperative anxiety risk predictors (30).

Regardless of a curative or a palliative approach, anxiety and depression rates in cancer patients are estimated at the level of 10% and 20%, respectively (31). The significant association of BC diagnosis and management with anxiety is validated by the reported depression and anxiety rate. In a recent meta-analysis, pooled postoperative depression and anxiety rates ranged at the levels of 4.7-78% and 12.5-71.3%, correspondingly (32). The long term duration of NMIBC treatment results to the development of post-traumatic stress disorder (PTSD) syndrome (33). In this cohort, it was confirmed that uncertainty and PTSD symptoms were inversely related to QoL as it was quantified by the EORTC QLQ-30 and QLQ-NMIBC24. Although post-resection anxiety and depression statuses are inferior compared to the general population, tumor and operative characteristics, catheterization time and morbidity rates significantly affect overall distress levels.

Our analyses showed a significant decrease of state anxiety, in contrast to the trait component, which remained stable. Both preoperative and postoperative STAI-Y scores were not associated to any of the studied characteristics. These are in accordance to the report of Palapattu et al. (34). In this study, general distress and anxiety decreased 1 month after surgery. Furthermore, demographic factors were not related to the overall distress prevalence. Moreover, Zhang et al., reported that NMIBC patients displayed more negative illness perceptions and these explained 42% and 39.5% of 3 and 12 months anxiety, respectively (35).

### Limitations

Prior to the appraisal of the results of the present study, several limitations should be acknowledged. First, the prospective methodology and the lack of patient and researcher blinding decreased the validity of our results. Although the statistical power of our findings was adequate, overall analyses would further benefit from a larger sample size. In addition to this, a heterogeneity was identified in terms of patient characteristics and habits, thus increasing the overall amount of bias. Finally, our long-term follow-up was defined by protocol at 12 months postoperatively. A longer period analysis would further elucidate the patient QoL and anxiety kinetics.

### Conclusions

In our study, an improvement in the Physical and Mental health aspect was identified during the follow-up period. Additionally, in contrast to Trait anxiety, State stress levels displayed a gradual declining trend. Preoperative Physical health was inversely correlated with age, while absence of alcohol was associated with lower Mental health. Overall, patient characteristics, habits and the administered treatment did not affect the postoperative QoL and anxiety. Due to several study limitations, larger scale studies with an extended follow up period are required for safe conclusions.

## Data Availability

The data are available by last author

## Disclosure

Conflict of Interest:

The authors declare that they have no conflict of interest.

## Supplementary Material

Supplementary Material Figures

## Notes

### Competing Interest Statement

The authors have declared no competing interest.

### Funding Statement

No funding

### Author Declarations

The study has been approved by the Ethics Committee of the University Hospital of Larissa, Greece

## References

1. Bray F, Ferlay J, Soerjomataram I, Siegel RL, Torre LA, Jemal A. Global cancer statistics 2018: GLOBOCAN estimates of incidence and mortality worldwide for 36 cancers in 185 countries. CA Cancer J Clin. 2018;68(6):394–424.

2. Richters A, Aben KKH, Kiemeney L. The global burden of urinary bladder cancer: an update. World J Urol. 2020;38(8):1895–904.

3. Yu EY, Nekeman D, Billingham LJ, James ND, Cheng KK, Bryan RT, et al. Health-related quality of life around the time of diagnosis in patients with bladder cancer. BJU Int. 2019;124(6):984–91.

4. Babjuk M, Burger M, Comperat EM, Gontero P, Mostafid AH, Palou J, et al. European Association of Urology Guidelines on Non-muscle-invasive Bladder Cancer (TaT1 and Carcinoma In Situ) - 2019 Update. Eur Urol. 2019;76(5):639–57.

5. Jung A, Nielsen ME, Crandell JL, Palmer MH, Smith SK, Bryant AL, et al. Health-related quality of life among non-muscle-invasive bladder cancer survivors: a population-based study. BJU Int. 2020;125(1):38–48.

6. Chung J, Kulkarni GS, Bender J, Breau RH, Guttman D, Maganti M, et al. Modifiable lifestyle behaviours impact the health-related quality of life of bladder cancer survivors. BJU Int. 2020;125(6):836–42.

7. Smith AB, Jaeger B, Pinheiro LC, Edwards LJ, Tan HJ, Nielsen ME, et al. Impact of bladder cancer on health-related quality of life. BJU Int. 2018;121(4):549–57.

8. Mohamed NE, Gilbert F, Lee CT, Sfakianos J, Knauer C, Mehrazin R, et al. Pursuing Quality in the Application of Bladder Cancer Quality of Life Research. Bladder Cancer. 2016;2(2):139–49.

9. Wettstein MS, Naimark D, Hermanns T, Herrera-Caceres JO, Ahmad A, Jewett MAS, et al. Required efficacy for novel therapies in BCG-unresponsive non-muscle invasive bladder cancer: Do current recommendations really reflect clinically meaningful outcomes? Cancer Med. 2020;9(10):3287–96.

10. Chen TH, Li L, Kochen MM. A systematic review: how to choose appropriate health-related quality of life (HRQOL) measures in routine general practice? J Zhejiang Univ Sci B. 2005;6(9):936–40.

11. Colombo R, Rocchini L, Suardi N, Benigni F, Colciago G, Bettiga A, et al. Neoadjuvant short-term intensive intravesical mitomycin C regimen compared with weekly schedule for low-grade recurrent non-muscle-invasive bladder cancer: preliminary results of a randomised phase 2 study. Eur Urol. 2012;62(5):797–802.

12. Morris K. Revising the Declaration of Helsinki. Lancet. 2013;381(9881):1889–90.

13. von Elm E, Altman DG, Egger M, Pocock SJ, Gotzsche PC, Vandenbroucke JP, et al. The Strengthening the Reporting of Observational Studies in Epidemiology (STROBE) statement: guidelines for reporting observational studies. J Clin Epidemiol. 2008;61(4):344–9.

14. Mitrakas LP, Zachos IV, Tzortzis VP, Gravas SA, Rouka EC, Dimitropoulos KI, et al. Previous Bladder Cancer History in Patients with High-Risk, Non-muscle-invasive Bladder Cancer Correlates with Recurrence and Progression: Implications of Natural History. Cancer Res Treat. 2015;47(3):495–500.

15. Pappa E, Kontodimopoulos N, Niakas D. Validating and norming of the Greek SF-36 Health Survey. Qual Life Res. 2005;14(5):1433–8.

16. Yoshimura K, Utsunomiya N, Ichioka K, Matsui Y, Terai A, Arai Y. Impact of superficial bladder cancer and transurethral resection on general health-related quality of life: an SF-36 survey. Urology. 2005;65(2):290–4.

17. Fountoulakis KN, Papadopoulou M, Kleanthous S, Papadopoulou A, Bizeli V, Nimatoudis I, et al. Reliability and psychometric properties of the Greek translation of the State-Trait Anxiety Inventory form Y: preliminary data. Ann Gen Psychiatry. 2006;5:2.

18. Sanli O, Dobruch J, Knowles MA, Burger M, Alemozaffar M, Nielsen ME, et al. Bladder cancer. Nat Rev Dis Primers. 2017;3:17022.

19. Sylvester RJ, Oosterlinck W, Witjes JA. The schedule and duration of intravesical chemotherapy in patients with non-muscle-invasive bladder cancer: a systematic review of the published results of randomized clinical trials. Eur Urol. 2008;53(4):709–19.

20. Sylvester RJ, Brausi MA, Kirkels WJ, Hoeltl W, Calais Da Silva F, Powell PH, et al. Long-term efficacy results of EORTC genito-urinary group randomized phase 3 study 30911 comparing intravesical instillations of epirubicin, bacillus Calmette-Guerin, and bacillus Calmette-Guerin plus isoniazid in patients with intermediate- and high-risk stage Ta T1 urothelial carcinoma of the bladder. Eur Urol. 2010;57(5):766–73.

21. Pereira MG, Lynch B, Hall-Faul M, Pedras S. Quality of life of women with urinary incontinence in rehabilitation treatment. J Health Psychol. 2019;24(2):254–63.

22. Moons P, Budts W, De Geest S. Critique on the conceptualisation of quality of life: a review and evaluation of different conceptual approaches. Int J Nurs Stud. 2006;43(7):891–901.

23. Cunillera O, Tresserras R, Rajmil L, Vilagut G, Brugulat P, Herdman M, et al. Discriminative capacity of the EQ-5D, SF-6D, and SF-12 as measures of health status in population health survey. Qual Life Res. 2010;19(6):853–64.

24. Singer S, Ziegler C, Schwalenberg T, Hinz A, Gotze H, Schulte T. Quality of life in patients with muscle invasive and non-muscle invasive bladder cancer. Support Care Cancer. 2013;21(5):1383–93.

25. Fung C, Pandya C, Guancial E, Noyes K, Sahasrabudhe DM, Messing EM, et al. Impact of bladder cancer on health related quality of life in 1,476 older Americans: a cross-sectional study. J Urol. 2014;192(3):690–5.

26. Schmidt S, Frances A, Lorente Garin JA, Juanpere N, Lloreta Trull J, Bonfill X, et al. Quality of life in patients with non-muscle-invasive bladder cancer: one-year results of a multicentre prospective cohort study. Urol Oncol. 2015;33(1):19 e7–e5.

27. Cox E, Saramago P, Kelly J, Porta N, Hall E, Tan WS, et al. Effects of Bladder Cancer on UK Healthcare Costs and Patient Health-Related Quality of Life: Evidence From the BOXIT Trial. Clinical Genitourinary Cancer. 2020;18(4):e418–e42.

28. Jadoon NA, Munir W, Shahzad MA, Choudhry ZS. Assessment of depression and anxiety in adult cancer outpatients: a cross-sectional study. BMC Cancer. 2010;10:594.

29. Erkilic E, Kesimci E, Soykut C, Doger C, Gumus T, Kanbak O. Factors associated with preoperative anxiety levels of Turkish surgical patients: from a single center in Ankara. Patient Prefer Adherence. 2017;11:291–6.

30. Kumar A, Dubey PK, Ranjan A. Assessment of Anxiety in Surgical Patients: An Observational Study. Anesth Essays Res. 2019;13(3):503–8.

31. Mitchell AJ, Chan M, Bhatti H, Halton M, Grassi L, Johansen C, et al. Prevalence of depression, anxiety, and adjustment disorder in oncological, haematological, and palliative-care settings: a meta-analysis of 94 interview-based studies. Lancet Oncol. 2011;12(2):160–74.

32. Vartolomei L, Ferro M, Mirone V, Shariat SF, Vartolomei MD. Systematic Review: Depression and Anxiety Prevalence in Bladder Cancer Patients. Bladder Cancer. 2018;4(3):319–26.

33. Jung A, Nielsen ME, Smith SK, Crandell J, Palmer MH, Bryant AL, et al. Uncertainty, post-traumatic stress, and quality of life in non-muscle-invasive bladder cancer survivors. Journal of Clinical Oncology. 2017;35(15_suppl):e16027–e.

34. Palapattu GS, Haisfield-Wolfe ME, Walker JM, BrintzenhofeSzoc K, Trock B, Zabora J, et al. Assessment of perioperative psychological distress in patients undergoing radical cystectomy for bladder cancer. J Urol. 2004;172(5 Pt 1):1814–7.

35. Zhang Z, Yang L, Xie D, Wang Y, Bi L, Zhang T, et al. Illness perceptions are a potential predictor of psychological distress in patients with non-muscle-invasive bladder cancer: a 12-month prospective, longitudinal, observational study. Psychology, Health & Medicine. 2019:1–11.

